# PC-POCS Sampler to reconstruct a sparse-view computer tomography image with both prior and measurements

**DOI:** 10.1101/2025.11.13.25340131

**Authors:** Yuchen Quan, Yaru Xue, Haisu Zhu

**Affiliations:** The College of Artificial Intelligence, China University of Petroleum, Beijing, Beijing 102249, China; MAIS & NLPR, Institute of Automation, Chinese Academy of Sciences, Beijing 100190, China

**Keywords:** Score-based generative model, Controllable generation, Projection to convex set, Filter back projection, Iterative reconstruction

## Abstract

Reconstructing medical images from partial measurements is a key inverse problem in computed tomography (CT), essential for reducing radiation exposure while maintaining diagnostic quality. Conventional supervised learning approaches depend on paired datasets generated under fixed acquisition settings, limiting their adaptability to unseen measurement processes. To overcome this, we propose a fully unsupervised framework based on score-based generative models. Our method learns the prior distribution of high-quality medical images and employs a physics-informed sampling strategy to reconstruct images consistent with both the learned prior and observed measurements. Experiments on multiple CT reconstruction tasks show that our approach achieves comparable or superior performance to existing methods while generalizing robustly across varying measurement conditions. The code is [available](https://anonymous.4open.science/r/PC-POCS-D2BE).

## 1 Introduction

With the rapid progress of machine learning, numerous methods have been developed for medical image reconstruction from limited measurements [1–6]. Most rely on supervised learning, where models are trained on large paired datasets of CT images and corresponding simulated measurements. However, these models depend on a fixed physical acquisition process—changes such as varying the number of CT projections require regenerating data and retraining, which is time-consuming and limits generalization. In some cases, performance may even degrade with more measurements [7].

To address these limitations, we propose an unsupervised framework that eliminates the need for paired data and fixed measurement settings. We model the prior distribution of medical images using a score-based generative model [8, 9], trained on high-quality images. During inference, a classifier-free [10] controllable sampling algorithm generates images consistent with both observed measurements and the learned prior, while accounting for the measurement physics. Once trained, the model can flexibly solve various linear inverse problems within the same image domain.

Extensive experiments on CT reconstruction tasks show that our method achieves comparable or superior results to existing unsupervised approaches [11,12]. More importantly, when acquisition conditions change (e.g., varying projection numbers), our method consistently outperforms all baselines. These results demonstrate its robustness, adaptability, and potential as a general-purpose framework for solving inverse problems in medical imaging.

## 2 Background

### 2.1 Variational Diffusion Models

In generative modeling, observed data are low-dimensional projections of latent variables encoding abstract features such as color, shape, and semantics. Like cave dwellers seeing only shadows, we cannot access the true sources directly but can infer their structure. Generative modeling thus aims to approximate these latent representations to understand and reconstruct data.

#### Hierarchical Variational Autoencoders

A Hierarchical Variational Autoen-coder (HVAE) [15,16] (see Figure 1) generalizes the VAE by introducing multiple latent hierarchies, where lower-level latents are generated from higher-level ones. While a general HVAE with T-level conditions each latent on all preceding latents, we focus on a Markovian HVAE (MHVAE), in which the generative process from a Markov chain: each latent *z*_*t*_ depends only on its immediate predecessor *z*_*t*+1_.

**Fig. 1.**
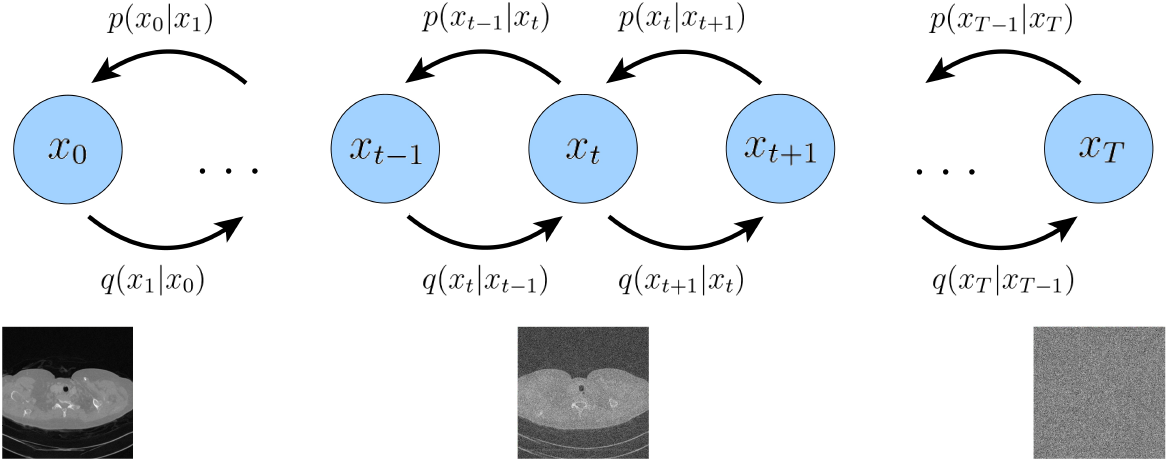
Score-based generative model showing the gradual recovery of a high-resolution CT image from noise

Mathematically, we represent the joint distribution and the posterior of a Markovian HVAE as:

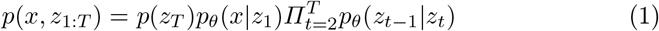

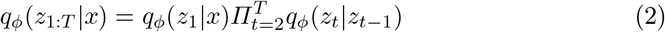

Substituting the original ELBO expression, we get the new objective:

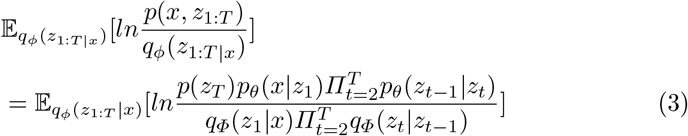

#### Unconditional Score-based Generative Mode

The Variational diffusion model [17–19] formulation has an equivalent Score-based Generative Modeling formulation. Arbitrary probability distributions can be written in the form:

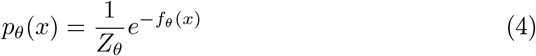

where *f*_*θ*_(*x*) is the energy function, often modeled by a neural network, and *Z*_*θ*_ is a normalizing constant to ensure that *p*_*θ*_(*x*)*dx* = 1. One way to avoid modeling the normalization constant is to use a neural network *s*_*θ*_(*x*) to learn the score function *∇lnp*(*x*) of distribution *p*(*x*) instead. This comes from taking the derivative of the ln of both sides of the equation above:

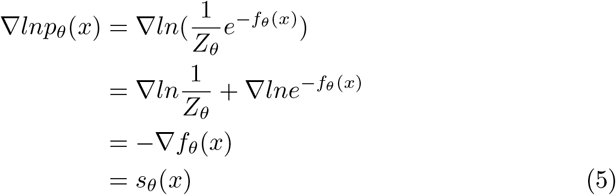

The equation above could be represented as a neural network without involving any normalization constants. The score model could be optimized by minimizing the Fisher Divergence with the ground truth score function:

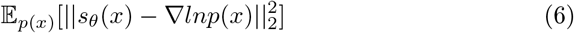

The score function defines a vector field over the entire space where the data *x* lies, pointing towards the modes. Then, by learning the score function of the true data distribution, we can generate samples by starting at an arbitrary point in the same space and iteratively following the score until a mode is reached. This sampling procedure is known as Langevin dynamics, and is mathematically described as:

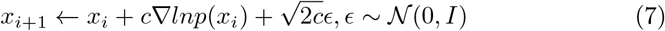

The initial value *x*_0_ is randomly sampled from a prior distribution. Eventually, we can choose a positive sequence of noise scale 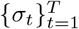 and then define a sequence of progressively perturbed data distributions:

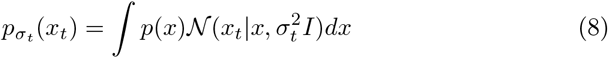

Then, a neural network *s*_*θ*_(*x, t*) is learned using denoising score matching to learn the score function for all noise levels:

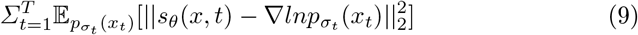

where *λ*(*t*) is a positive weighting function depending on the noise level *t*. Song [9] introduces annealed Langevin Dynamics sampling (Figure 2a), where initialization comes from a handcrafted prior, and each step starts from the previous final samples. This resembles the Markovian HVAE interpretation of a Variational Diffusion Model, where a randomly initialized data vector is iteratively refined over decreasing noise levels.

**Fig. 2.**
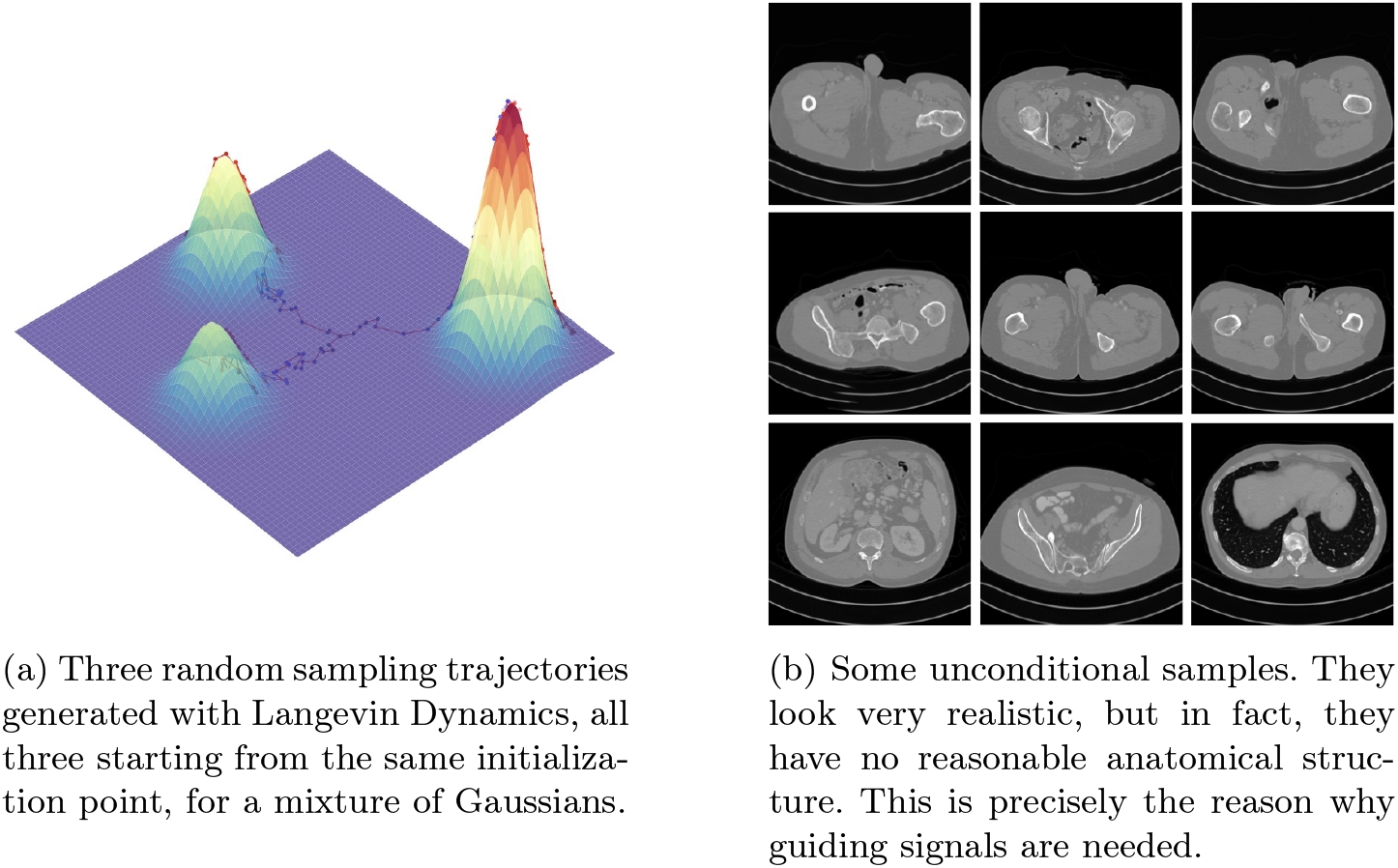
The sampling trajectories and some unconditional samples.

With infinitely many noise scales, image perturbation can be modeled as a stochastic process governed by an SDE. Sampling reverses this SDE, requiring score estimation at each noise level. Different SDE parameterizations define distinct perturbation schemes; here, we use the Variance Exploding (VE) SDE (see Figure 2b).

#### Classifier-Free Guidance

Learning two separate diffusion models is expensive; in this work, we just consider the classifier-free guidance [10, 20, 21]. To derive the score function under Classifier-Free Guidance, first, we have:

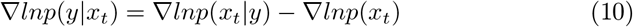

Then, substituting this into the *classifier-guidance* equation *∇lnp*(*x*_*t*_|*y*) = *∇lnp*(*x*_*t*_) + *γ∇lnp*(*y*|*x*_*t*_), get:

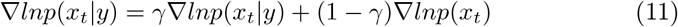

The first term is the conditional score, and the second is the unconditional score. The coefficient *γ* controls conditioning influence: *γ* = 0 ignores it (unconditional), *γ* = 1 learns the standard conditional distribution, and *γ >* 1 emphasizes the conditional score while moving away from the unconditional one.

### 2.2 Sparse-view Computer Tomography Inverse Problems

An inverse problem seeks to recover an unknown signal from observed measurements. Let **x** ∈ ℛ^*n*^ be the signal and **y** ∈ ℛ^*m*^ the measurements modeled as **y** = **Ax** + *ϵ*, where **A** ∈ ℛ ^*m*\**n*^ and *ϵ* is noise. When *m < n*, the problem is ill-posed without prior knowledge. Assuming **x** ~ *p*(**x**), the likelihood is *p*(**y**|**x**) = *q*_*ϵ*_(**y** − **Ax**). The inverse problem is then solved by inferring the posterior *p*(**x**|**y**) and sampling from it.

## 3 Methodology

### 3.1 Projection Onto Convex Sets (POCS)

In mathematics, the projection onto convex sets (POCS) method, also known as the alternating projection method, finds a point in the intersection of two closed convex sets. When the sets are affine spaces, the iterates converge—if the intersection is non-empty—not just to any point in the intersection but specifically to the **orthogonal** projection of the point onto it. POCS always has the following iteration form:

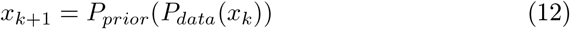

where *P*_*data*_ denotes projecting data *x* onto the data fidelity set, and *P*_*prior*_ denotes projecting data *x* onto the prior set.

### 3.2 Imputation in the Radon domain

Imputation is a special case of conditional sampling. Let the known and unknown dimensions of **y** be Ω(**y**) and 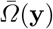, and let 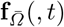 and 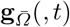 denote the drift and diffusion coefficients restricted to the unknown dimensions. Fow VE SDEs, **f** (, *t*) is element-wise and **g**(, *t*) is diagonal. Thus, 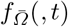 applies the element-wise drift only to unknown dimensions, and 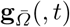 is the sub-matrix of the diffusion restricted to them.

In this setting, *P*_*prior*_ specifically denotes the samples transformed by the forward model from the diffusion model at each timestep, and *P*_*prior*_ denotes the constraint coming from the forward model. Because the Radon transform operator **A** is not orthogonal, we need an orthogonal term (**AA**^*T*^)^*†*^ analogous to the filtering step to implement the orthogonal projection, to make POCS work well.

### 3.3 PC-POCS sampling

Combining POCS with the PC sampling method, we derive the PC-POCS algorithm. Specifically, we use the discretized timestep, from *t* = 1 to *t* = *ϵ*, and each timestep conducts two steps of update. The first step is used to predict the diffusion trajectory:

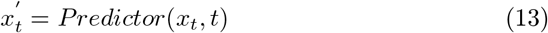

which always has the form of

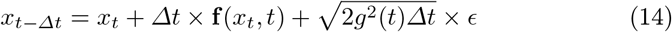

The second step is used to correct the trajectory. The first part of the second step is Langevin correct:

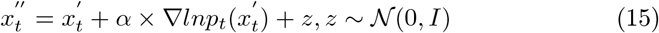

After the corrector, come into the Kaczmarz iteration to satisfy the projection consistency:

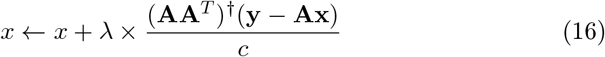

where **A** denotes the forward radon projection operator, **A**^*T*^ denotes the back projection operator, (**AA**^*T*^)^*†*^ denotes the filter back projection operator. **y** denotes the measurement, i.e., sparse-view sinogram, and *λ* denotes the stepsize scheduler coefficient, *c* = **A**^*T*^ (**A**(*I*)) denotes the normalization constant. The details of the whole algorithm are in the pseudocode.

#### Algorithm 1

PC-POCS

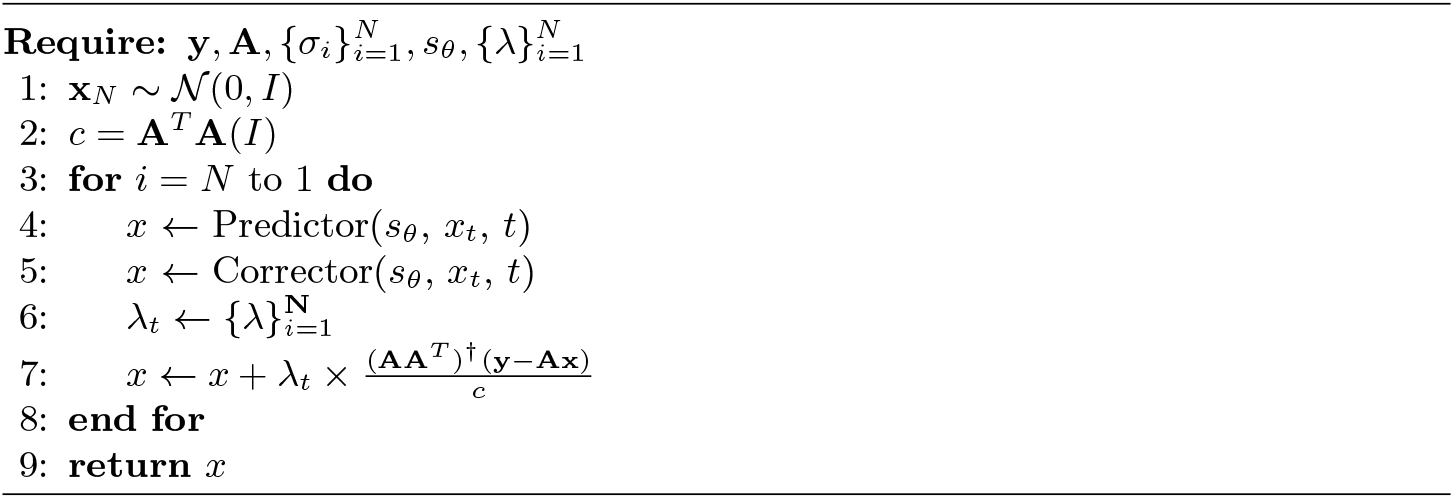

## 4 Experiment

For all tasks, we aim to verify the superiority of our method against other diffusion model-based approaches.

### 4.1 Dataset

We use the dataset from the AAPM 2016 CT Low-Dose Grand Challenge, acquired in a fan-beam geometry with varying parameters. Data preprocessing follows the procedure in [22]. The helical cone-beam projections are converted to fan-beam geometry via single-slice rebinning, and reconstructed using standard filtered backprojection (FBP) to obtain axial images of size 512 *×* 512. These slices are then resized to 512 *×* 512 and used to train the score function. The dataset comprises 9 volumes (3142 slices) for training and 1 volume (823 slices) for testing. To simulate sparse-view measurements, we retrospectively apply a parallel-beam geometry for simplicity.

### 4.2 Network training

For the CT score function, we train the ncsnpp network with setting *λ* = *σ*^2^(*t*), and *ϵ* = 10^−5^. Training is conducted by using two RTX 3080 Ti for 100 epochs, and the average loss eventually converges to 2 *×* 10^−2^.

### 4.3 Evaluation metrics

We use three standard metrics: SSIM, PSNR, and FSIM for quantitative evaluation. FSIM may be a relatively novel metric, and makes an improvement on the SSIM, introducing phase consistency with position as a variable and weighting the SSIM based on this. The basic formula of FSIM is:

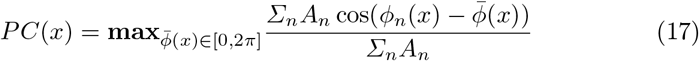

The equation above expresses the image signal as a Fourier series expansion, where each component has a corresponding phase. 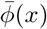 represents the average phase of all components, *ϕ*_*n*_(*x*) denotes the phase of each individual component, and *A*_*n*_ is the amplitude.

### 4.4 Comparative experiment with the State-of-the-Art Methods

To the best of our knowledge, [11] and [12] are the only two methods that solve CT reconstruction directly with diffusion models. So, we compare our method against theirs. We call the method from [11] as Song, and the approach from [12] as MCG. To control the variables, the experiments of the three methods were all conducted under the condition of sparsity of 6 (i.e., the number of projections was 30). The numerical results of the experiment are shown in TABLE. 1, and the visualization results are shown in Figure 3

**Fig. 3.**
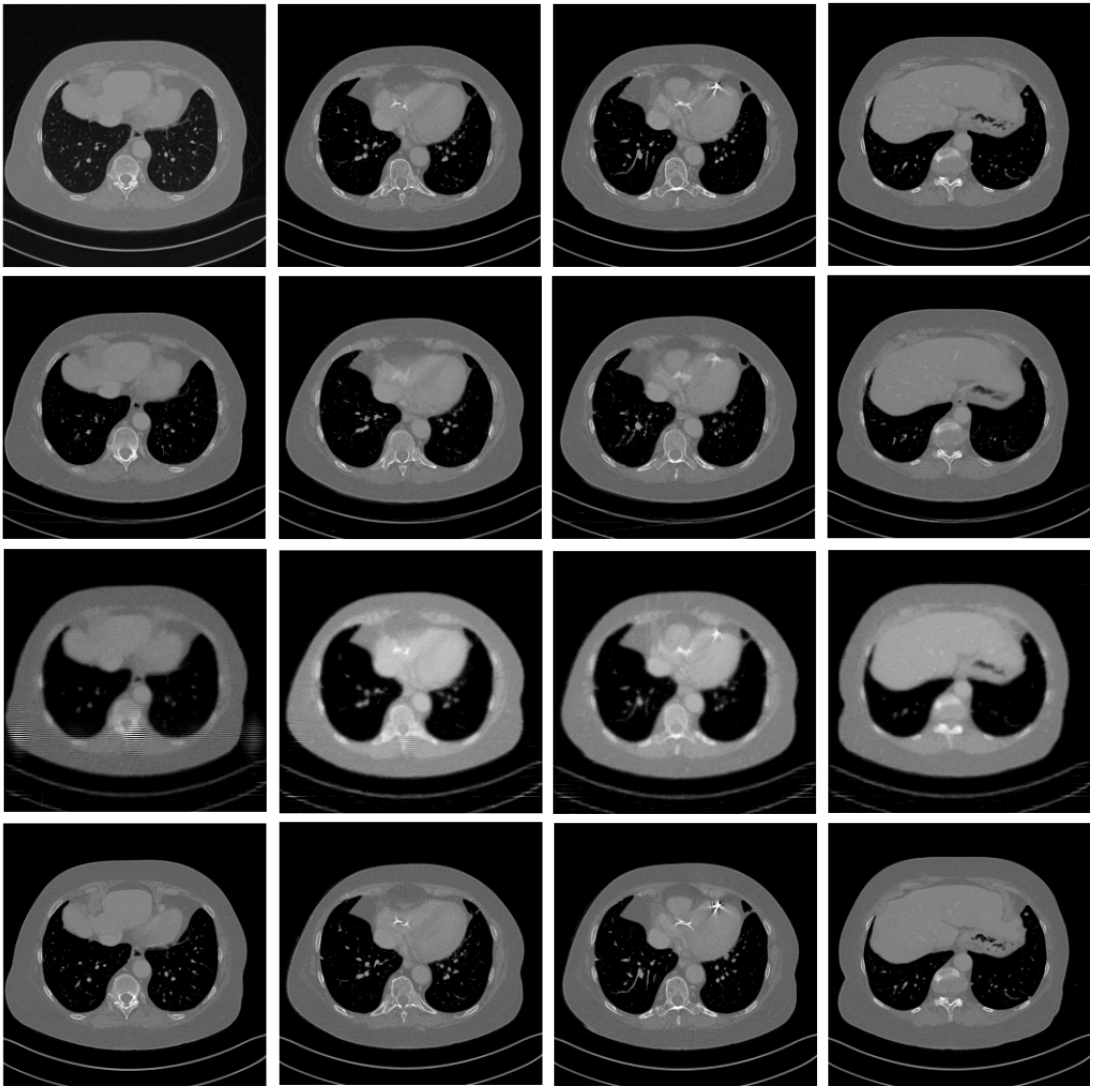
The reconstruction results of all three methods. From top to bottom, the corresponding are PC-POCS(Proposed), MCG, Song, and the ground truth. PC-POCS has the best performance. Zoom in to see details.

From Table 1, the PC-POCS method outperforms the other two methods across all three metrics, achieving either the highest or second-highest values among all slices of patient L096. Visually, under a sparsity level of 6 (30 projections), PC-POCS provides the best reconstruction quality.

**Table 1.** Quantitative results of different methods on AAPM dataset. We experiment on the slice index of 268 ~ 287. Due to space limitations, we have only presented some of the random results. The **Bold** characters represent our method. Best numerical results are **red**; second best are **blue**.

| Slice<br>Index | MCG |  |  | Song |  |  | PC-POCS |  |  |
| --- | --- | --- | --- | --- | --- | --- | --- | --- | --- |
|  | SSIM | FSIM | PSNR | SSIM | FSIM | PSNR | SSIM | FSIM | PSNR |
| 268 | 0.8245 | 0.8948 | 28.0463 | 0.7534 | 0.8494 | 19.3313 | 0.8902 | 0.9544 | 33.7343 |
| 269 | 0.8185 | 0.8927 | 27.9395 | 0.7598 | 0.8506 | 19.0324 | **0.8952** | 0.9559 | 34.0099 |
| 270 | 0.8217 | 0.8935 | 28.1502 | 0.7601 | 0.8489 | 19.8642 | 0.8915 | 0.9550 | 33.7837 |
| 271 | 0.8265 | 0.9006 | 28.5476 | 0.7555 | 0.8466 | 19.2071 | 0.8886 | 0.9524 | 33.8067 |
| 272 | 0.8193 | 0.8939 | 27.9205 | 0.7509 | 0.8447 | 19.4014 | 0.8882 | 0.9526 | 33.6799 |
| 273 | 0.8229 | 0.8978 | 28.2543 | 0.7569 | 0.8447 | 21.0755 | 0.8909 | 0.9531 | 33.6392 |
| 274 | 0.8124 | 0.8886 | 27.4984 | 0.7603 | 0.8446 | 21.1059 | 0.8867 | 0.9529 | 33.4561 |
| 275 | 0.8168 | 0.8909 | 27.5838 | 0.7659 | 0.8447 | 23.4483 | 0.8880 | 0.9539 | 33.4843 |
| 276 | 0.8198 | 0.8914 | 27.4239 | 0.7627 | 0.8461 | 24.8852 | 0.8914 | 0.9544 | 33.4510 |
| 277 | 0.8141 | 0.8901 | 27.2613 | 0.7693 | 0.8460 | 25.2738 | 0.8868 | 0.9522 | 33.0534 |
| 278 | 0.8255 | 0.8977 | 27.8815 | 0.7695 | 0.8424 | 24.8828 | 0.8862 | 0.9530 | 33.3379 |
| 279 | 0.8114 | 0.8894 | 27.2112 | 0.7661 | 0.8481 | 21.6348 | 0.8853 | 0.9530 | 33.3544 |
| 280 | 0.8231 | 0.9005 | 28.0072 | **0.7706** | 0.8499 | 21.5181 | 0.8867 | 0.9544 | 33.5495 |
| 281 | 0.8228 | 0.9012 | 27.9217 | 0.7138 | 0.8355 | 23.2376 | 0.8921 | 0.9559 | 33.8156 |
| 282 | 0.8216 | 0.9021 | 28.4271 | 0.7609 | 0.8478 | 22.7803 | 0.8876 | 0.9558 | 33.7879 |
| 283 | 0.8263 | 0.9069 | 28.5272 | 0.7701 | 0.8483 | 23.2087 | 0.8881 | 0.9553 | 33.8860 |
| 284 | 0.8239 | 0.9066 | 28.5368 | 0.7629 | **0.8517** | **25.7797** | 0.8889 | 0.9558 | 33.9643 |
| 285 | 0.8197 | 0.9007 | 28.3875 | 0.7656 | **0.8554** | 23.0487 | 0.8930 | 0.9567 | 34.2841 |
| 286 | **0.8274** | **0.9095** | **28.9279** | 0.7659 | 0.7865 | **25.6019** | 0.8927 | **0.9596** | **34.5028** |
| 287 | **0.8316** | **0.9145** | **29.2608** | **0.7707** | 0.8478 | 21.7916 | **0.8950** | **0.9614** | **34.6276** |

### 4.5 Ablation Study

Determining the lower bound of sparsity is a crucial task for different reconstruction methods. In general, higher sparsity leads to poorer reconstruction quality. For example, when using SSIM as the evaluation criterion, a reliably reconstructed medical image typically exhibits an SSIM value greater than 0.7. Images reconstructed below this threshold usually suffer from severe structural loss and therefore lack clinical value.

Due to this consideration, we conducted an ablation experiment to determine the sparsity corresponding to the minimum trusted image. The relationship between the sparsity and the number of projections is:

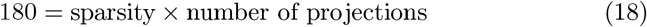

From Fig. 5, the relationship between the sparsity (number of projections) and metrics is obvious. For the integrity of the experiment, we also conducted experiments on FSIM and PSNR, and provided the results. According to Fig. 5, when sparsity = 26, the SSIM reaches the lower bound of 0.7.

For the sake of demonstrating the performance of our method, we conduct an ablation study on the other two methods. The experiment result is also shown in Fig. 5. Besides that, the visualization results of this ablation study are also provided in Figure 4. Our proposed method needs less physical information than the other two methods and has the highest threshold sparsity for reconstruction.

**Fig. 4.**
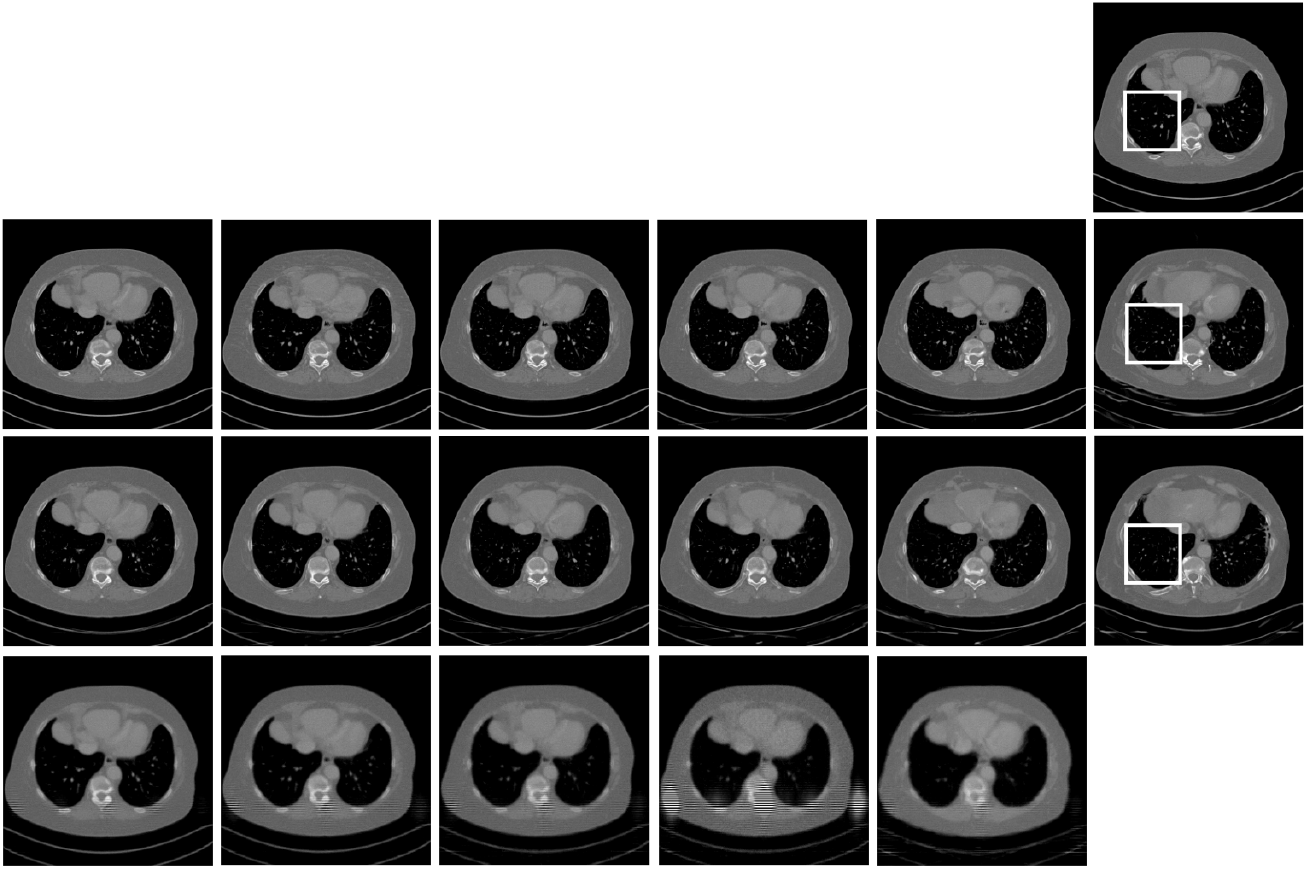
The ablation study result of slice 290 of patient L096. From left to right, the images correspond to different sparsity levels. From upwards to downwards, the images correspond to ground truth, PC-POCS, MCG, and Song, respectively. For PC-POCS, the sparsity is set to 6, 8, 10, 12, 18, 30. Because the quality of the image is very low under the method of Song, we just set the sparsity of 2, 3, 5, 6, 10 for reference. We find that under high sparsity, some subtle structure is lost, for example, the part in **the white border** is different in the ground truth and the reconstruction under high sparsity. You can zoom in to see more details of the image.

**Fig. 5.**
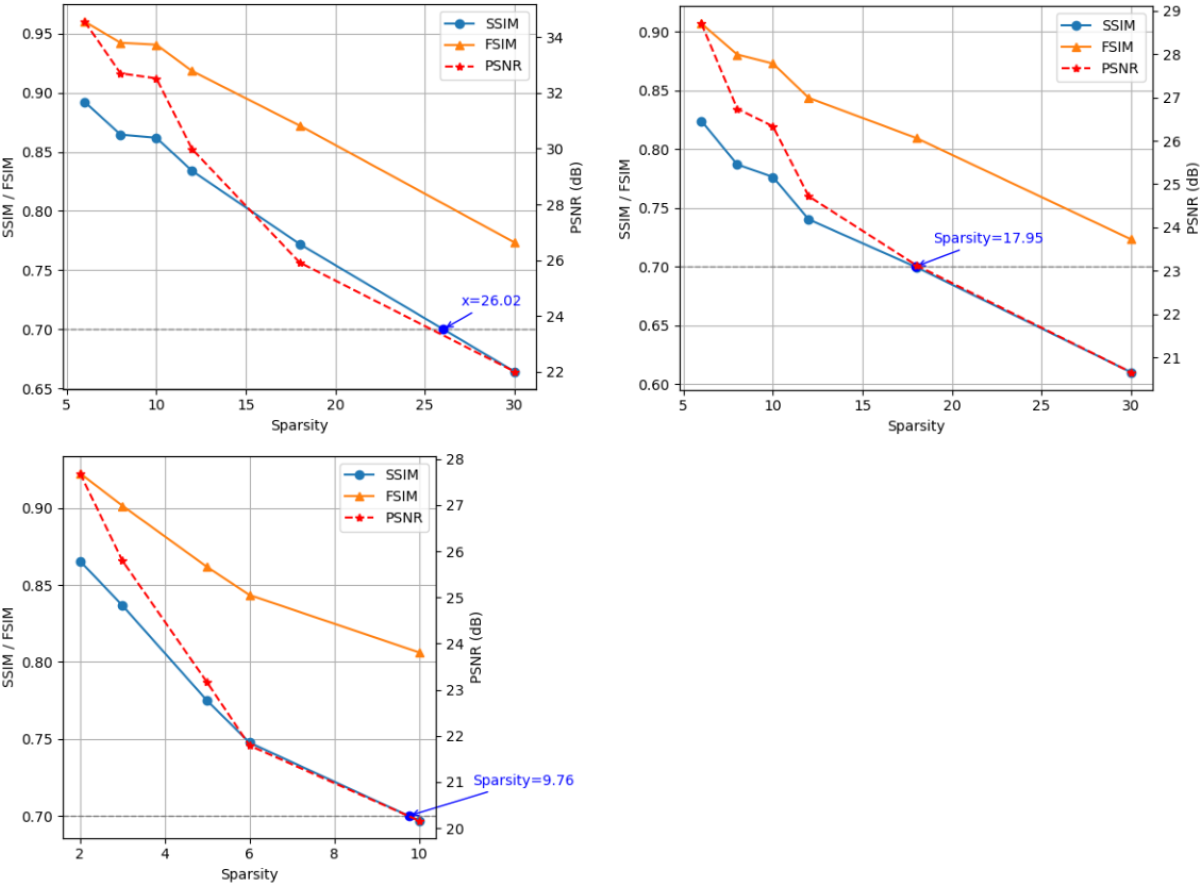
The relationship between the sparsity and each metric. With the increase of sparsity, the number of projections decreases, so the true physical information decreases, and the quality of the reconstruction images deteriorates. From left to right, and from top to bottom, the results are PC-POCS, MCG, and Song, respectively. According to the result image, when sparsity = 26, 18, 10, the SSIM reaches the threshold of 0.7, respectively. So the proposed sampler PC-POCS has the best effect according to the analysis above.

## 5 Conclusion

In this work, we propose a general framework that significantly enhances the solver performance of score-based generative models for low-dose CT reconstruction. Our method outperforms the current state-of-the-art approaches. Moreover, ablation studies demonstrate that it achieves reliable reconstruction under high sparsity and exhibits stronger robustness than the other two methods.

## Data Availability

All data produced in the present work are contained in the manuscript.

https://www.cancerimagingarchive.net/collection/ldct-and-projection-data/

